# Post-infection depressive, anxiety and post-traumatic stress symptoms: a retrospective cohort study with mild COVID-19 patients

**DOI:** 10.1101/2020.08.25.20182113

**Authors:** Flavia Ismael, João C. S. Bizario, Tatiane Battagin, Beatriz Zaramella, Fabio E. Leal, Julio Torales, Antonio Ventriglio, Megan E. Marziali, Silvia S. Martins, João M. Castaldelli-Maia

## Abstract

**Background:** It remains unclear whether COVID-19 is associated with psychiatric symptoms during or after the acute illness phase. Being affected by the disease exposes the individual to an uncertain prognosis and a state of quarantine. These factors can predispose individuals to the development of mental symptoms during or after the acute phase of the disease. There is a need for prospective studies assessing mental health symptoms in COVID-19 patients in the post-infection period.

**Methods:** In this retrospective cohort study, nasopharyngeal swabs for COVID-19 tests were collected at patients’ homes under the supervision of trained healthcare personnel. Patients who tested positive for COVID-19 and were classified as mild cases (N=895) at treatment intake were further assessed for the presence of mental health disorders (on average, 56.6 days after the intake). We investigated the association between the number of COVID-19 symptoms at intake and depressive, anxiety and post-traumatic symptoms, adjusting for previous mental health status, time between baseline and outcome, and other confounders. Multivariate logistic regression and generalized linear models were employed for categorical and continuous outcomes, respectively.

**Outcomes:** A clinically significant level of depressive, anxiety and post-traumatic stress symptoms were reported by 26.2% (N=235), 22.4% (N=201), and 17.3% (N=155) of the sample. Reporting an increased number of COVID-related symptoms was associated with clinically significant level of depressive (aOR=1.059;95%CI=1.002-1.119), anxiety (aOR=1.072;95%CI=1.012-1.134), and post-traumatic stress (aOR=1.092;95%CI=1.024-1.166) symptoms. Sensitivity analyses supported findings for both continuous and categorical measures.

**Interpretation:** Exposure to an increased number of COVID-19 symptoms may predispose individuals to depressive, anxiety and post-traumatic symptoms after the acute phase of the disease. These patients should be monitored for the development of mental health disorders after COVID-19 treatment discharge. Early interventions, such as brief interventions of psychoeducation on coping strategies, could benefit these individuals.

**Funding:** The city health department of São Caetano do Sul (*Secretaria Municipal de Saúde da Prefeitura de São Caetano do Sul*) funded the establishment and implementation of the COVID-19 platform.

## 1. Introduction

The COVID-19 pandemic has affected a significant amount of individuals worldwide(1). Despite the efforts to limit viral spread, cases are increasing worldwide and deaths are continually occurring(2). This pandemic is generating further mental issues such as insomnia, anxiety, depression, stress, anger, and fear(3). Those directly or indirectly affected by the virus could be more disturbed by these symptoms(3,4). Word cloud studies indicate that uncertainties about lack of COVID-19 tests and medical supplies are common(5).

There is still much uncertainty about the best treatment to be administered to individuals affected by the disease(5). Though highly transmissible, most cases present with mild symptoms(2). However, having been affected by the disease exposes the individual to an uncertain prognosis and a need to quarantine to mitigate viral spread(6). These factors can predispose individuals to the development of mental symptoms during or after the acute phase of the disease. It is unclear whether COVID-19 can produce psychiatric symptoms during or after the acute illness phase(6,7).

In general, survivors of critical illnesses have a high level of mental symptoms after the condition improves. Depression, anxiety and post-traumatic stress disorder (PTSD) are among the most reported events in patients with these conditions(8). Patients infected with SARS-CoV-1 had a high rate of depressive symptoms during follow-up after the acute phase of the disease(9-11). These symptoms lasted for an extended period, being reported up to a year after the improvement in SARS-CoV-1 symptoms(11). Anxiety symptoms were also reported during the post-SARS-CoV-1 follow-up(9,10).

Some studies in Asia investigated depression and/or anxiety in patients admitted in hospitals due to COVID-19(12-15). In a case-control design, Guo et al.(12) investigated the mental status and inflammatory markers of 103 COVID-19 hospitalized mild patients, matching them with controls that were COVID-19 negative. Hu et al.(13) carried out a cross-sectional survey with COVID-19 inpatients in two isolation wards of a COVID-19 designated hospital. Zhang et al.(15) evaluated the prevalence and severity of depression and anxiety within patients recently recovered from COVID-19 infection, who were under quarantine. In Vietnam, Nguyen et al.(14) carried out a cross-sectional study with individuals infected by COVID-19 attending outpatient departments of nine hospitals and health centers across the country. All these studies found increased levels of both anxiety and depression (6.8-21.0% and 7.4-31.5%, respectively). There was no follow-up study to investigate prospective symptoms of depression and anxiety in COVID-19 patients.

The ongoing COVID-19 pandemic has disrupted the lives of many across the globe, resulting in an increased burden of physical and mental health consequences. Through this analysis, we investigated the association between COVID-19 symptoms and post-infection depressive, anxiety and post-traumatic symptoms among a sample of patients diagnosed with mild COVID-19 in Brazil. There is a need for prospective studies assessing mental health symptoms in COVID-19 patients, evaluating the post-infection period in other regions of the world.

## 2. Methods

### 2.1. Ethical Approval

The present study was approved by the local ethics committee (*Comissão de Ética para Análise de Projeto de Pesquisa* - CAPPesq, protocol No. 32293020.9.0000.5510, approved on July 13^th^, 2020).

### 2.2. Study Design

This was a retrospective cohort study. All people who tested positive for COVID-19 and classified as mild cases at treatment intake in the public health system of a Brazilian city with around 160,000 inhabitants (baseline: April 6^th^ to July 15^th^) were considered for the presence of mental health disorders in a follow-up online assessment (outcome: July 20^th^ to early August 7^th^). We investigated the association between the number of COVID-19 symptoms at intake and clinically significant levels of depressive, anxiety and post-traumatic stress symptoms in the follow-up assessment, adjusting for previous mental health status, and the time between the baseline and outcome, among other possible confounders. Sensitivity analyses were carried out where we excluded: (i) individuals with a short time between baseline and outcome assessment (up to 14 days), because these individuals could be in the late active phase of the COVID-19 disease, and (ii) those who progressed to a more severe case of COVID-19.

### 2.3. Sample

Residents of the municipality ≥18 years of age with suspected COVID-19 symptoms were encouraged to contact an specific website/phone platform for assessing COVID-19 (access at https://coronasaocaetano.org/) (baseline: April 6^th^ to July 15^th^). They were invited to complete an initial screening questionnaire that included socio-demographic data; information on symptoms type, onset and duration; and recent contacts. People meeting the suspected COVID-19 case definition (i.e., having at least two of the following symptoms: fever, cough, sore throat, coryza, or change in/loss of smell (anosmia); or one of these symptoms plus at least two other symptoms consistent with COVID-19) were further evaluated, whilst people not meeting these criteria were reassured, advised to stay at home and contact the service again if they were to develop new symptoms or the worsening of current ones. Patients were then asked by a medical student to complete a risk assessment. There were no refusals. All pregnant women, and patients meeting pre-defined triage criteria for severe disease, were advised to attend a hospital service - either an emergency department or outpatient service, depending on availability.

All other patients were offered a home visit for self-collection of a nasopharyngeal swab (NPS – both nostrils and throat), which were collected at the patients’ homes under the supervision of trained healthcare personnel. More details can be found in Leal et al.(16). Due to shortages of some reagents, two RT-PCR platforms were used at different times during the study: ALTONA RealStar® SARS-CoV-2 RT-PCR Kit 1.0 (Hamburg, Germany) and the Mico BioMed RT-qPCR kit (Seongnam, South Korea). For serology, we tested 10μL of serum or plasma (equivalent in performance) using a qualitative rapid chromatographic immunoassay (Wondfo Biotech Co., Guangzhou, China), that jointly detects anti-SARS172 CoV-2 IgG/IgM. The assay has been found to have a sensitivity of 81.5% and specificity of 99.1% in a U.S. study (16). In our local validation, after two weeks of symptoms, the sensitivity in RT-PCR confirmed cases (N=59) was 94.9%, and specificity in biobank samples (N=106) from 2019 was 100% (16). Patients testing RT-PCR negative were followed up by the primary health care program of their residential area. They were advised to contact the platform for additional consultation if they developed new symptoms.

COVID-19 positive patients presenting dyspnea, tachypnea, persistent fever (≥ 72 hours), or mental health change at any timepoint were further evaluated by a doctor, who could refer the patient to healthcare services (i.e., moderate and severe cases). All the other patients who tested positive were classified as mild and followed-up by phone (N=1,757). These mild patients were invited to participate in the present retrospective cohort study, in which we assessed depressive, anxiety, and post-traumatic stress symptoms (outcome: July 20^th^ to early August 7^th^). We had a response rate of 50.9%. Table S1 presents differences a comparison between those that agreed to participate (N=895) and those that did not (N=862). People that agreed to participate in the study were younger and reported more headaches, anosmia and dysgeusia, and less tachypnea and joint pain than those that refused to be part of the study. More importantly, no significant difference was found regarding the total number of COVID-19 symptoms, which was our main exposure measure.

### 2.4. Measures

All the exposure measures were collected online via the dedicated Corona São Caetano web platform (access at https://coronasaocaetano.org/) or by phone. The outcomes were assessed online only.

#### 2.4.1. Exposure (COVID-19 symptoms)

Patients testing positive for COVID-19 via RT-PCR were followed up to 14 days (a maximum of 7 phone calls) from completion of their initial questionnaire. They were contacted every 48 hours by a medical student (supervised by a medical doctor) who completed another risk assessment and recorded any ongoing or new symptoms. Following the COVID-19 clinical assessment protocol of São Caetano do Sul(16), the following COVID-19 symptoms were assessed during these contacts: dyspnea; tachypnea; persistent fever (≥ 72 hours); mental health disturbance (e.g., changes in consciousness, thought, perception); fever (at any timepoint); cough; sore throat; nasal congestion; coryza; headache; fatigue; asthenia; lack of appetite; myalgia; joint pain; diarrhea; nausea; vomit; anosmia; and dysgeusia. The total number of symptoms during the treatment was the primary exposure investigated in the present study.

#### 2.4.2. Outcomes (Mental Health Symptoms)

The GAD-7 scale is an instrument for assessing, diagnosing and monitoring anxiety symptoms. It was created by Spitzer et al.(17). It was validated by Kroenke et al.(18), according to the criteria of the Diagnostic and Statistical Manual of Mental Disorders – Fourth Edition (DSM-IV), for the assessment of signs and symptoms of anxiety disorder, and also to classify severity levels. This study uses the Brazilian Portuguese validated version(19). GAD-7 consists of seven items, on a four-point scale: 0 (not at all), 1 (several days), 2 (more than half the days), and 3 (nearly every day). The total score ranges from 0 to 21, assessing the frequency of signs and symptoms of anxiety over a two-week period. No missingness was observed in any of the question items. A cutoff ≥10 was used for defining a clinically significant level of anxiety symptoms (20). In our sample, we found a Cronbach’s alpha of 0.92 (Table S1).

The PHQ-9 scale is an adaptation of the PRIME-MD (21). It is a brief instrument for assessing, diagnosing and monitoring depressive symptoms. It was validated by Spitzer et al.(22) and by Kroenke et al.(23). The present study uses a version which has been translated and validated to Brazilian Portuguese(24). PHQ-9 was created based on the DSV-IV criteria for Major Depressive Disorder, for the assessment of its signs and symptoms, and also to classify severity levels. It consists of nine items, arranged on a frequency four-point scale: 0 (not at all), 1 (several days), 2 (more than half the days), and 3 (nearly every day). Its score ranges from 0 to 21, assessing the frequency of signs and symptoms of anxiety over two weeks. No missingness was observed in any of the question items. A cutoff ≥10 was used for defining a clinically significant level of depressive symptoms(25). In our sample, we found a Cronbach’s alpha of 0.90 (Table S1).

Weathers et al.(26) developed the PCL-C scale, which was translated, adapted and validated to Brazilian Portuguese(27,28) to assess the consequences of different types of traumatic experiences. It is based on the DSM-III diagnostic criteria for PTSD. The patient must report the levels of last-month disturbance by 17 items, using a severity scale ranging from 1 (not at all), 2 (a little bit), 3 (moderately, 4 (quite a bit), and 5 (extremely). No missingness was observed in any of the question items. A cutoff ≥44 for defining a clinically significant level of post-traumatic stress symptoms(29). In our sample, we found a Cronbach’s alpha of 0.94 (Table S1).

#### 2.4.3. Possible confounders

Lifetime diagnosis of psychiatric disorder (yes vs. no), current psychiatric treatment (yes vs. no), age (continuous: 18-88 years), gender (male vs. female), education (up to high school vs. more than high school), civil status (married vs. single, which included previously married), income level (as defined by the Brazilian Institute of Geography and Statistics: up to three times the typical salary for a minimum wage job vs. more), current health treatment for any acute or chronic medical condition (yes vs. no) and time between the treatment intake and mental assessment (continuous: 6-116 days), were assessed as potential confounders.

### 2.5. Statistical Analysis

STATA software version 16.2 was used to run the analysis. Initially, we performed a comparison between those who attended the mental health follow-up assessment and were included in the present study (N= 895) and those who did not, using logistic regression models. This comparison was performed to identify any potential baseline difference between the groups, which could generate bias to our outcome analysis (e.g., higher number of COVID-19-related symptoms among those not included). Our final analytical sample included 895 participants. We first conducted a descriptive analysis of the COVID-19 treatment intake profile, sociodemographic measures, and the health profile of included patients. Secondly, we described the mean and prevalences of clinically significant level of anxiety, depressive and post-traumatic stress symptoms in these patients. We then created scatterplot figures for continuous outcomes across time. Multivariate logistic regression models for categorical outcomes (binarized scales) were carried out. These models were adjusted for all aforementioned confounders listed in section 2.4.3. Two distinct models were carried out, one which included lifetime psychiatric diagnosis, and the other included current psychiatric treatment, due to significant correlation between these two variables determined via pairwise testing (p<0.05). We subsequently ran sensitivity analyses, where we excluded: (i) individuals with a short time between baseline and outcome assessment, as individuals could be in the late active phase of the COVID-19 disease (≧14 days), (ii) those who progressed to a more severe COVID-19 case, and (iii) those with a previous psychiatric diagnosis. In a final sensitivity analysis, we ran multivariate generalized linear models (GLM) for the continuous outcomes. Based on a previous study(30), gamma-family GLM with log link were the models of choice, because of a log-normal distribution of the continuous outcomes of depression, anxiety and PTSD in our sample (Figures S1, S2, and S3).

## 3. Results

Table 1 shows descriptive analysis of our sample (N=895). The majority were female (60.4%), married (51.4%), and had up to high-school education (60.4%) and three minimum salaries per month of income (58.9%). Around one in every five individuals have had a psychiatric disorder during lifetime (20.1%). Only about half of these individuals have been undergoing psychiatric treatment (10.5%). Current health treatment was reported by 43.1% of the sample. Regarding COVID-19 symptomatic profile, patients had a mean of 4.2 COVID-19-related symptoms. The most common symptoms were anosmia (51.9%), dysgeusia (49.6%), cough (43.1%), headache (41.3%), and fatigue (36.9%), being reported by more than 35% of the sample.

**Table 1.** Descriptive analysis of 895 patients classified as having mild COVID-19 at treatment intake, São Caetano do Sul, 2020.

|  |  |  |
| --- | --- | --- |
|  | Mean/n | SE/% |
| **Sociodemographic** |  |  |
| *Age* | 40.79 | 0.45 |
| *Female gender* | 541 | 60.44 |
| *Married* | 460 | 51.40 |
| *Education (up to high-school)* | 541 | 60.44 |
| *Montly income (up to 3 minimum salaries)* | 536 | 59.89 |
| **Health profile** |  |  |
| *Current health treatment* | 386 | 43.13 |
| *Lifetime psychiatric diagnosis* | 180 | 20.11 |
| *Current psychiatric treatment* | 95 | 10.53 |
| **COVID-19 profile** |  |  |
| *Number of symptoms* | 4.19 | 0.10 |
| *Dyspnea\** | 11 | 1.25 |
| *Tachypnea\** | 3 | 0.74 |
| *Persistent fever\** | 4 | 0.56 |
| *Mental health change\** | 2 | 0.23 |
| *Fever\*\** | 65 | 7.40 |
| *Cough\*\** | 379 | 43.12 |
| *Sore throat\*\** | 159 | 18.09 |
| *Nasal congestion\*\** | 277 | 31.55 |
| *Coryza\*\** | 213 | 24.32 |
| *Headache\*\** | 363 | 41.39 |
| *Fatigue\*\** | 324 | 36.94 |
| *Asthenia\*\** | 179 | 20.41 |
| *Lack of appetite\*\** | 230 | 26.32 |
| *Myalgia\*\** | 259 | 29.57 |
| *Joint pain\*\** | 84 | 9.58 |
| *Diarrhea\*\** | 105 | 11.97 |
| *Nausea\*\** | 129 | 14.69 |
| *Vomit\*\** | 21 | 2.39 |
| *Anosmia\** | 456 | 51.94 |
| *Dysgeusia\** | 435 | 49.60 |
| *\*Assessed by a healthcare professional* |  |  |
| *\*\*Self-reported* |  |  |

Table 2 presents depressive, anxiety and post-traumatic stress symptoms and disorders in the sample. Clinically significant levels of depressive, anxiety and post-traumatic stress symptoms were reported by 26.2% (N = 235), 22.4% (N = 201), and 17.3% (N = 155) of the sample. Among these patients, 39.2% (N = 92), 37.8% (N = 76), and 50.3% (N = 78), had a previous psychiatric diagnosis during lifetime. On average, we assessed patient mental health almost two months after the treatment intake (mean = 56.6 days, 95%CI = 54.7-58.5), with the vast being assessed after the acute phase of the disease (78.7%, N = 840). Few patients (6.7%, N = 61) were referred for in-person consultation.

**Table 2.** Depressive, anxiety and post-traumatic stress symptoms and disorders among 895 patients who had previously mild COVID-19, São Caetano do Sul, 2020.

|  |  |  |  |  |  |
| --- | --- | --- | --- | --- | --- |
|  | Mean | 95%CI | Cutoff | n | % |
| *Depressive Symptoms / Depression (PHQ-9)* | 6.65 | 6.24-7.06 | $\geq 10$ | 235 | 26.26 |
| *Anxiety symptoms / Anxiety Disorder (GAD-7)* | 5.97 | 5.61-6.33 | $\geq 10$ | 201 | 22.46 |
| *Post-traumatic stress symptoms / PTSD (PCL-C)* | 31.58 | 30.72-32.45 | $\geq 44$ | 155 | 17.32 |
| *Time of Mental Health Assessment (days after intake)* | 56.61 | 54.71-58.51 | $\geq 14$ | 840 | 78.73 |
| *Severity (referred to in-person medical consultation)* | N.A. | N.A. | N.A. | 61 | 6.78 |

Figures 1A, 1B, and 1C present linear prediction graphs with 95% confidence intervals of continuous outcomes (x-axis) by the number of baseline COVID-19 symptoms (y-axis). The latter predicted increasing scores of depressive, anxiety, and post-traumatic stress symptoms. Figures S4, S5, and S6 present scatterplots of scores of depression, anxiety, and post-traumatic stress (y-axis) by the time of the mental health assessment (x-axis). There were wide ranges of scores for all the outcomes, more concentrated in the lower severity levels during the entire period (from 1 week to almost four months). For all the outcomes, a similar pattern of distribution was found through the time of the mental health assessment.

**Figure 1A-1B-1C.**
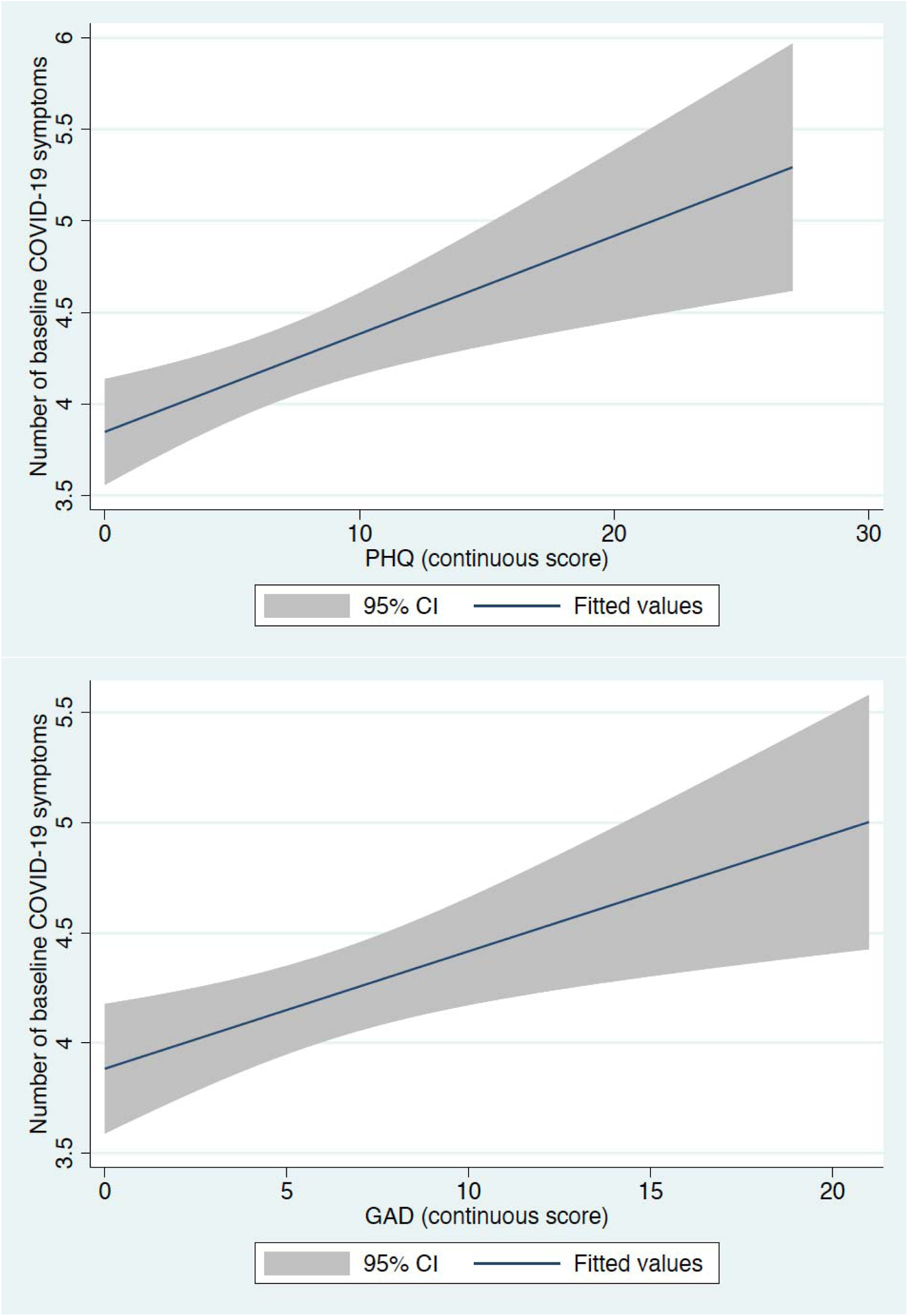
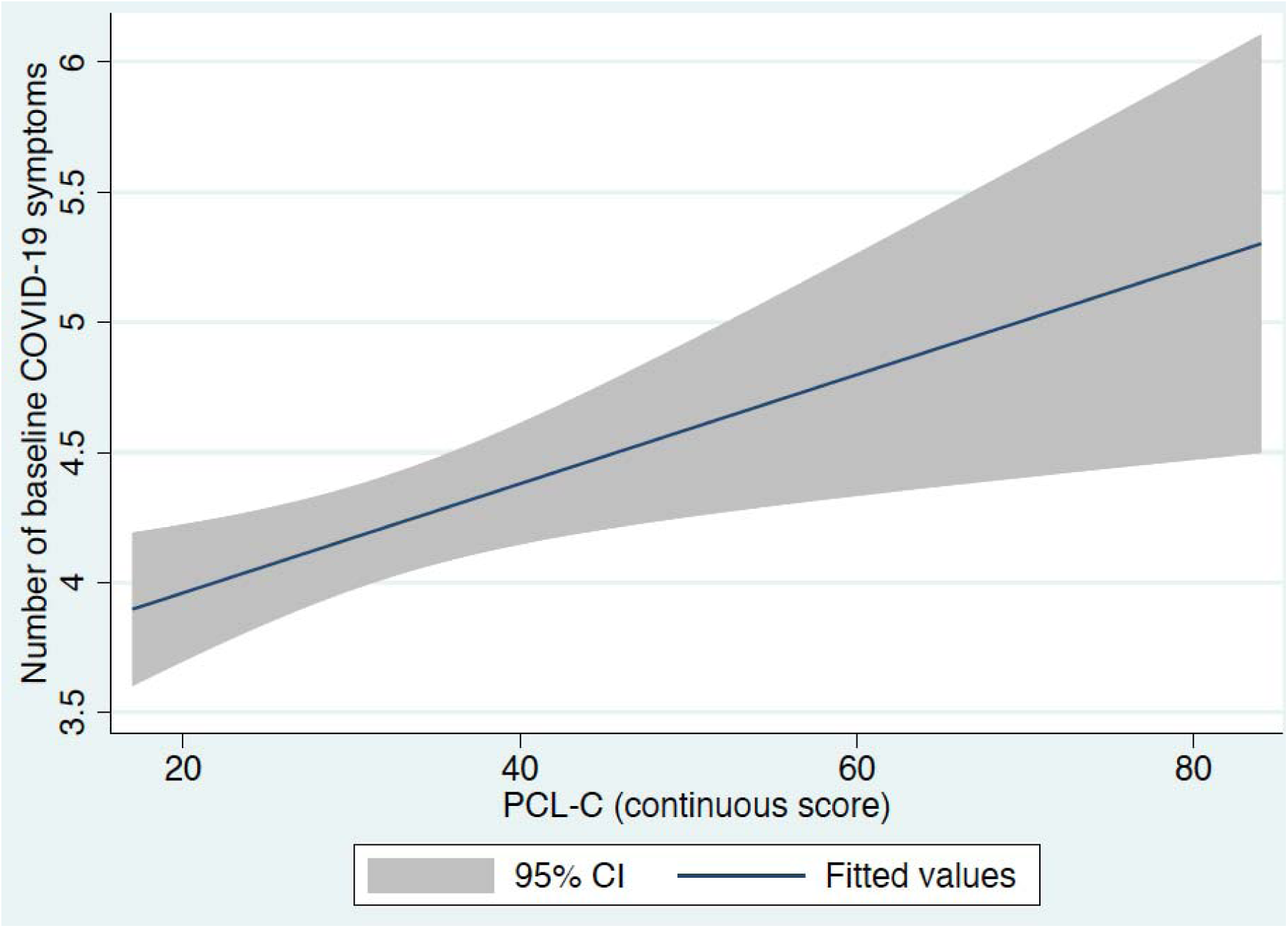
Linear prediction graphs with 95% confidence intervals of continuous outcomes (x-axis) by the number of baseline COVID-19 symptoms (y-axis).

Table 3 presents the results of the logistic regression models of the exposure (previous total number of symptoms of COVID-19) for the categorical outcomes (clinically significant level of depressive, anxiety and post-traumatic stress symptoms). The exposure was significantly associated with all the outcomes, after adjustment for all confounders.

**Table 3.**
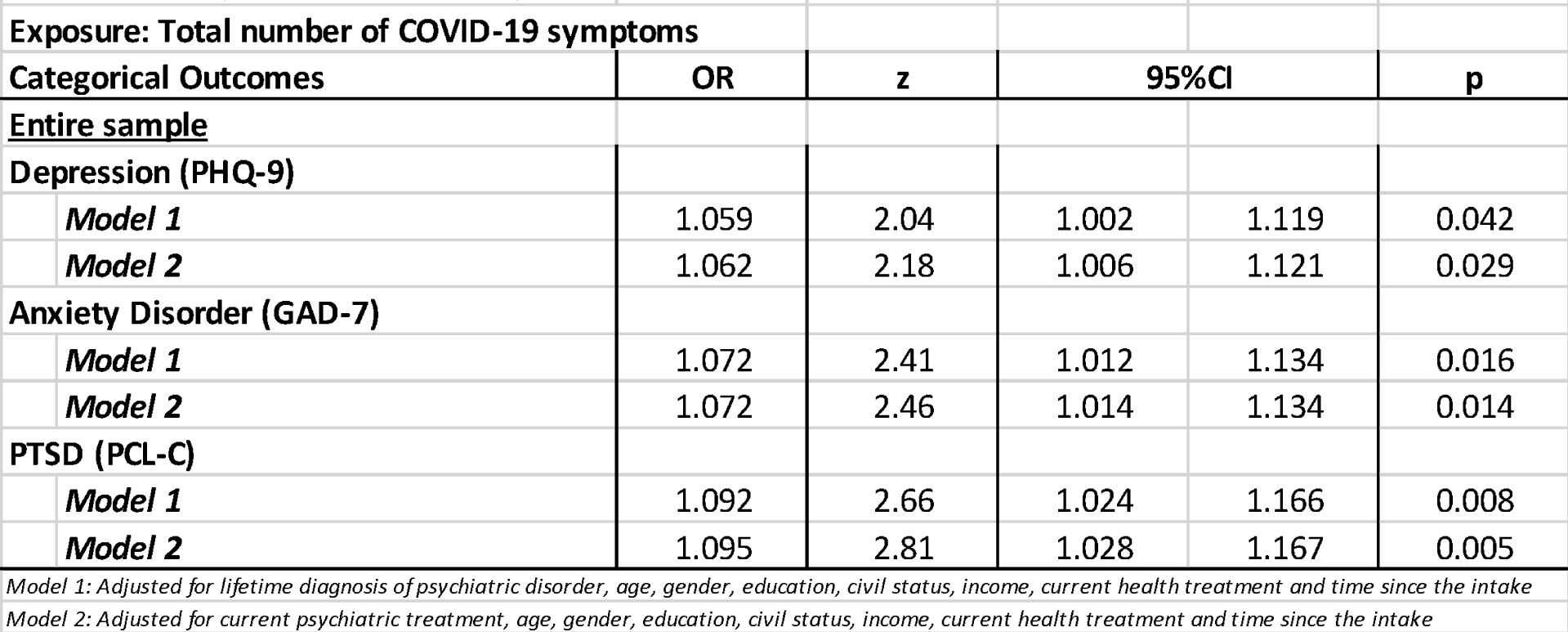
Results of the multivariate logistic regression models among 895 patients who had previously mild COVID-19, São Caetano do Sul, 2020.

In the sensitivity analysis (Table 4), these results remained significant after the exclusion of (i) individuals with a short time between baseline and outcome assessment (≥ 14 days), as individuals could be in the late active phase of the COVID-19 disease, (ii) those who progressed to a more severe COVID-19 case, and (iii) those with a previous psychiatric diagnosis. In the final sensitivity analysis (GLM for continuous outcomes), we found a significant relationship between number of COVID-19 symptoms and all the outcomes, with the exception of post-traumatic stress symptoms when adjusting for lifetime psychiatric disorder (p = 0.053).

**Table 4.**
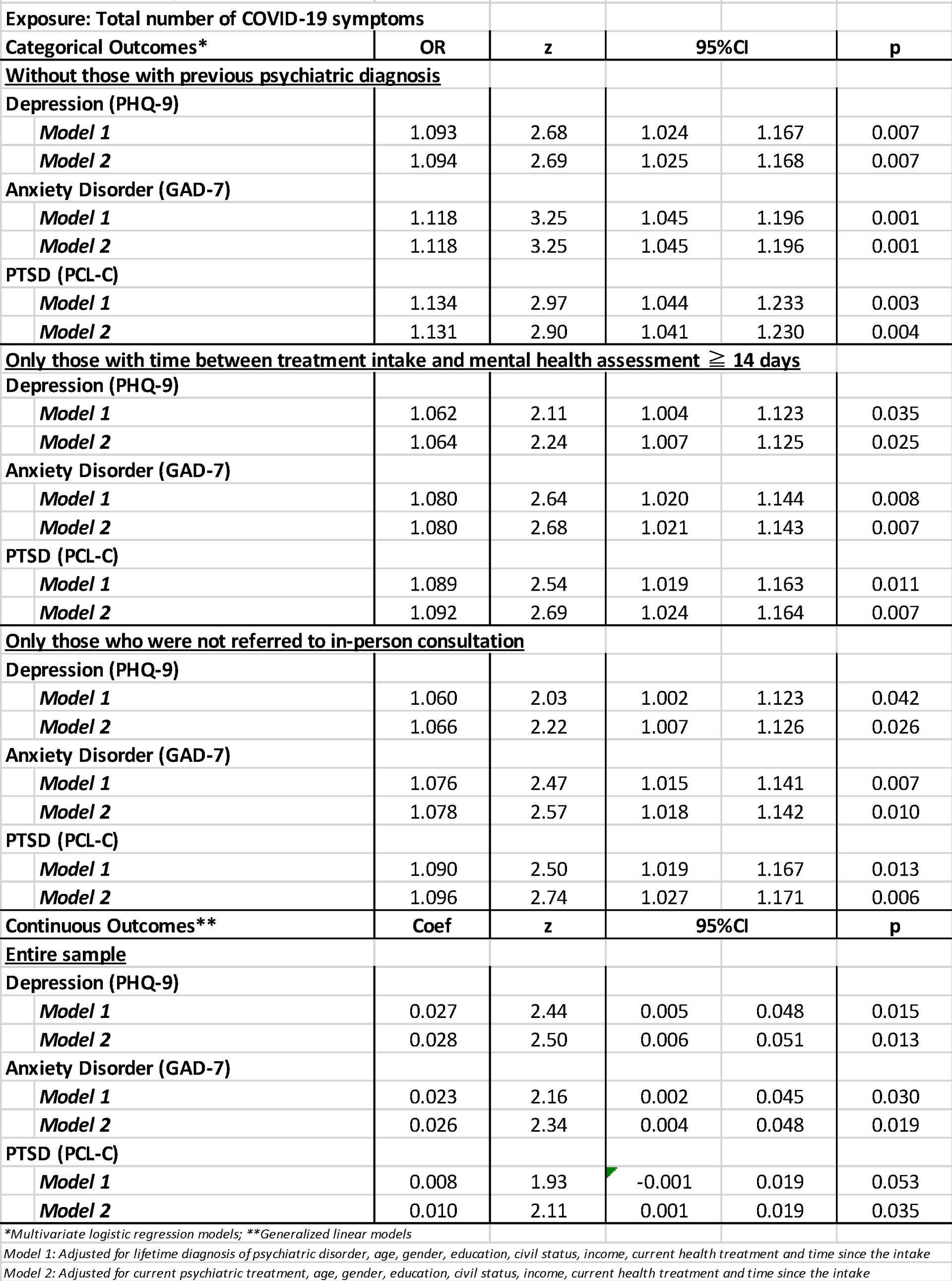
Results of the sensitivity analysis among 895 patients who had previously mild COVID-19, São Caetano do Sul, 2020.

## 4. Discussion

The present study aimed to examine the post-infection levels of mental health disorders among individuals with mild COVID-19 disease. We aimed to investigate whether COVID-19 infection symptomatology could be associated with mental health disorders. We found that an increased number of COVID-related symptoms were associated with a clinically significant level of depressive, anxiety, and post-traumatic stress symptoms. Sensitivity analyses supported those findings for the categorical clinical diagnosis of such disorders. More importantly, our findings adjusted for confounders that could increase the vulnerability of mental health disorders. These results shed light on a significant subpopulation at risk for mental disorders. This has been the largest study evaluating mental health symptoms in patients who had COVID-19 disease to date, and the only study assessing mental health status of patients with prior COVID-19 infections.

Four studies in Asia investigated depressive and/or anxiety symptoms in COVID-19 patients using the same scales used in the present study(12-15). Prevalence of depression and anxiety varied between 7.4-31.5%.and 6.8-21.0%, respectively(12-15). All of these studies were conducted in Asia (three in China and one in Vietnam). The prevalence of a clinically significant level of depressive symptoms in our study (26.2%) is included within this interval, but clinically significant anxiety symptoms level was greater (22.4%) than previously reported values (6.8-21.0%). Our results were more similar to those found by Zhang et al (15), who sampled home-quarantined COVID-19 patients. The lowest depression and anxiety prevalences were found in the Guo et al.(12) study, which included COVID-19 hospitalized patients.

A clinically significant level of post-traumatic stress symptoms, reported by 17.3% (N=155) of respondents with mild COVID-19 in our study, has remained largely unassessed within the general population during the COVID-19 pandemic. Research regarding post-traumatic stress symptoms, using the PCL-C scale, has been predominantly carried out within specified populations; within China, 16.3% of nurses in the Hubei province(31), 2.9% of university students(32) and 14.4% of youth(33) reported post-traumatic stress symptoms. Among a sample in Spain, some of whom experienced COVID-19 symptoms, 15.8% reported post-traumatic stress symptoms(34): a similar prevalence to that observed within this sample. Further, research conducted regarding the SARS outbreak in 2003 has demonstrated that 13-21.7% of healthcare workers experienced post-traumatic stress symptoms(35). Previous estimates of post-traumatic stress levels within Brazil were 8.5%(36) demonstrating that the prevalence within individuals presenting with mild COVID-19 is increased in comparison to past estimates.

Our results support the hypothesis that the prevalence of clinically significant levels of depressive, anxiety and post-traumatic stress symptoms were elevated in people with increased number of COVID-19 symptoms at baseline. These findings echo warnings from the previous SARS outbreak, wherein survivors of SARS infections experienced increased psychological distress, persisting one year or more subsequent to the outbreak(11). Similar findings were observed following the occurrence of the Middle East Respiratory Syndrome Coronavirus (MERS-CoV) in 2015, indicating that survivors experienced mental health consequences following the outbreak(37). Mental health supports should be strengthened, and healthcare systems must prepare for an influx of individuals experiencing psychological distress as a result of the COVID-19 pandemic. Following the PTSD model, these individuals should be referred to early interventions. Brief interventions of psychoeducation on coping strategies have been effective in promoting mental health among individuals who experienced traumatic life events(38). Internet-based psychological intervention for acute COVID-19 patients has also been described, and could be an interesting early-intervention tool for those who experience psychological distress during this phase(39).

It is unclear whether COVID-19 can produce psychiatric symptoms during or after the acute illness phase(4). Neuropsychiatric issues, such as: headaches, paresthesia, myalgia, impaired consciousness, confusion or *delirium*, and cerebrovascular diseases have been reported among individuals with COVID-19(7). However, the symptoms assessed in the present study (i.e., depressive, anxiety and post-traumatic stress) are substantially different from neuropsychiatric symptoms observed among some individuals in the acute phase of COVID-19. In addition, we found no differences of level of mental health symptomatology depending on the time of assessment after the acute phase of the disease. Thus, it seems improbable that depressive, anxiety and post-traumatic stress symptoms could be a direct effect of the COVID-19 infection. Rather, it is likely that the increased prevalence of mental health disorders post-COVID-19 is resultant from the psychosocial context of the pandemic(40). People who have been infected with COVID-19 have likely experienced long periods of quarantine, and some have reported fear of transmitting the virus to members of their social and familial networks(41). This, in combination with uncertainties surrounding treatment and clinical course(12), could be working synergistically to worsen mental health symptoms. Future studies should explore neurobiological effects of SARS-Coronavirus-2 and mental health impacts.

### 4.1. Strengths and Limitations

Assessing people for depressive, anxiety, and post-traumatic stress symptoms at different timepoints should be noted as an important limitation of the present study. However, we adjusted all the logistic regression and GLM models to the time of assessment and also conducted sensitivity analyses, excluding those who could potentially be assessed during the acute phase of COVID-19 and testing whether the continuous or categorical version. We were also not able to assess other important behavioral disorders (i.e., substance use and sleep disorders). However, we were able to assess the most prevalent disorders following traumatic experiences in almost a thousand COVID-19 patients through reliable measures both for exposure and outcomes, with an acceptable response rate. The patients included in the present study were slightly different from those who did not attend the invitation. Despite the latter being older, no significant difference was found for the total number of COVID-19 symptoms, which was our exposure measure. The main issue for generalization of our findings was the inclusion of individuals dependent on the public healthcare sector only.

### 4.2. Conclusion

Exposure to increased levels of COVID-19 symptomatology may predispose individuals to clinically significant levels of depressive, anxiety and post-traumatic stress symptoms after the acute phase of the disease, independently of previous psychiatric diagnosis. These patients should be monitored for the development of mental health disorders after COVID-19 treatment discharge. Early mental health intervention such as psychotherapy and supportive groups could play an important role in preventing incident mental health problems in these people. It is probable that the increased prevalence of mental health disorders post-COVID-19 is due to the social and psychological context of the disease. However, further studies should investigate the possible neurobiological mechanisms linking COVID-19 and mental health conditions.

## Data Availability

All the data associated with this paper has been included in the results and supplementary files.

---

**Table S1.** Results of the scales test (Cronbach’s alpha).

| **Table S1.** Results of the scales test (Cronbach's alpha). |  |  |  |
| --- | --- | --- | --- |
| **Scale** | **PHQ-9** | **GAD-7** | **PCL-C** |
| *Average interitem covariance* | 0.43 | 0.56 | 0.57 |
| *Number of items in the scale* | 9 | 7 | 17 |
| *Scale reliability coefficient* | 0.90 | 0.92 | 0.94 |

**Figures S1-S2-S3.**
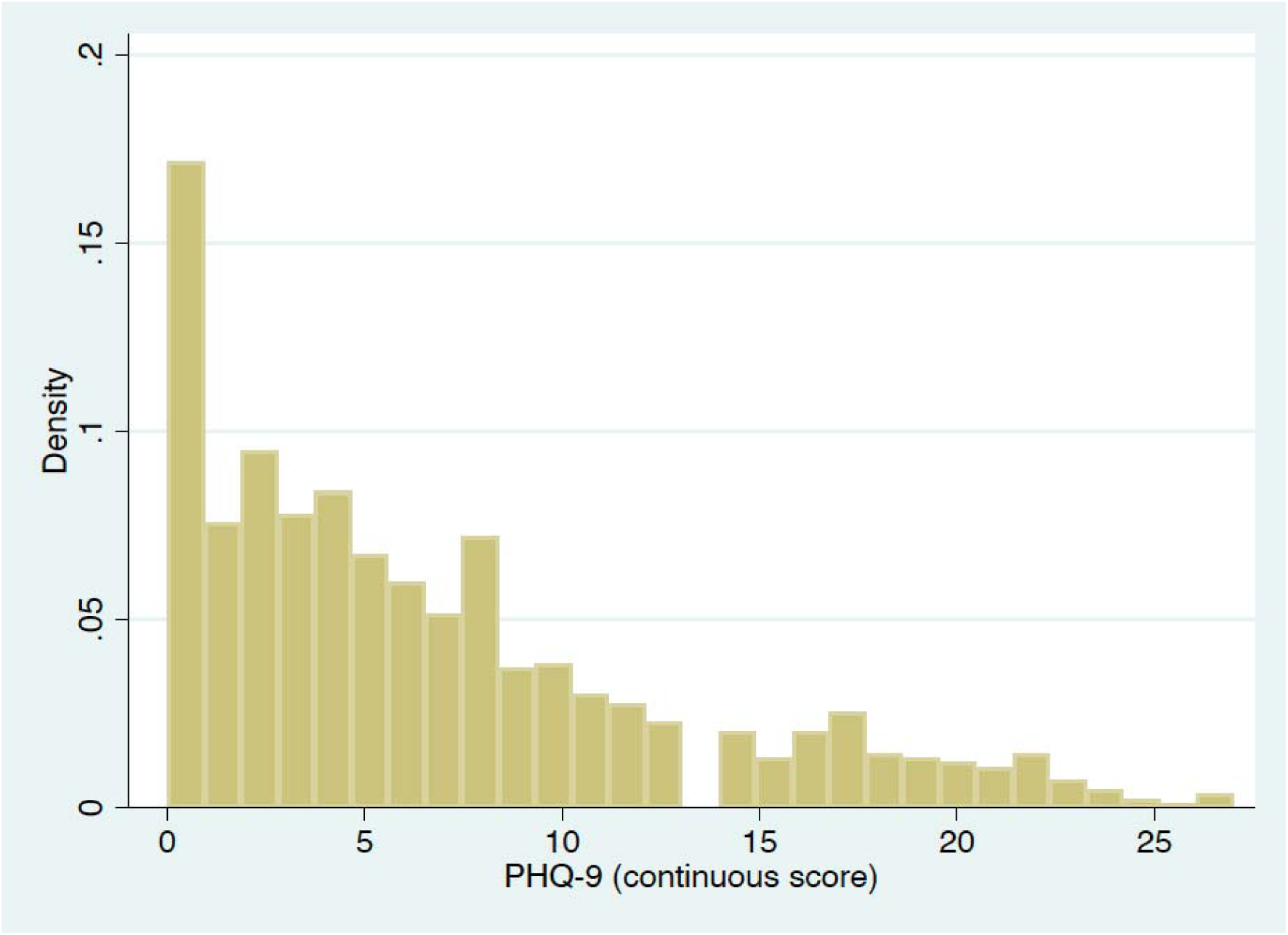
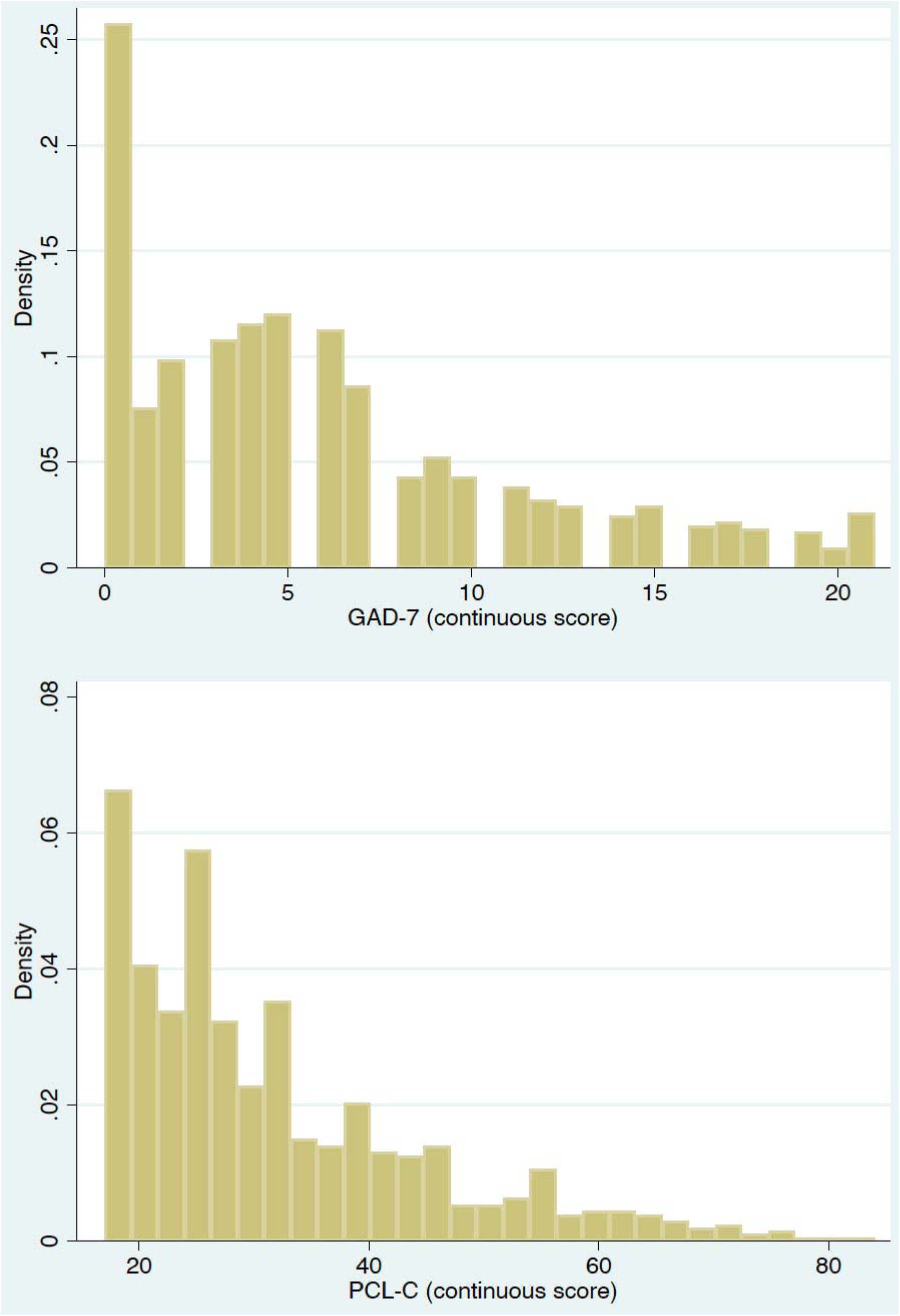
Distribution of the continuous outcomes of depression, anxiety and PTSD in our sample.

**Table S2.** Logistic regression models for follow-up versus missing among those classified as having mild COVID-19 patients at treatment intake, São Caetano do Sul, 2020.

|  | **OR** | **z** | **95%CI** |  | **p** |
| --- | --- | --- | --- | --- | --- |
| *Age* | 0.98 | -6.12 | 0.98 | 0.99 | <0.001 |
| *Positive Test Day* | 1.00 | 0.72 | 1.00 | 1.00 | 0.474 |
| *Number of symptoms* | 1.02 | 1.41 | 0.99 | 1.06 | 0.158 |
| *Breathless\** | 0.52 | -1.64 | 0.24 | 1.14 | 0.101 |
| *Tachypnea\** | 0.26 | -2.09 | 0.07 | 0.92 | 0.037 |
| *Persistent fever\** | 0.47 | -1.22 | 0.14 | 1.57 | 0.221 |
| *Mental health change\** | 0.47 | -0.86 | 0.09 | 2.59 | 0.387 |
| *Fever\*\** | 0.87 | -0.78 | 0.61 | 1.24 | 0.434 |
| *Cough\*\** | 1.05 | 0.48 | 0.86 | 1.27 | 0.634 |
| *Sore throat\*\** | 0.99 | 0.12 | 0.77 | 1.26 | 0.907 |
| *Nasal congestion\*\** | 1.03 | 0.30 | 0.84 | 1.27 | 0.763 |
| *Coryza\*\** | 1.09 | 0.76 | 0.87 | 1.37 | 0.449 |
| *Headache\*\** | 1.35 | 2.99 | 1.11 | 1.65 | 0.003 |
| *Fatigue\*\** | 1.13 | 1.24 | 0.93 | 1.38 | 0.215 |
| *Asthenia\*\** | 0.83 | -1.63 | 0.66 | 1.04 | 0.104 |
| *Anorexia\*\** | 0.99 | 0.11 | 0.80 | 1.23 | 0.912 |
| *Myalgia\*\** | 0.96 | -0.34 | 0.78 | 1.19 | 0.731 |
| *Joint pain\*\** | 0.68 | -2.48 | 0.51 | 0.92 | 0.013 |
| *Diarrhea\*\** | 1.32 | 1.77 | 0.97 | 1.80 | 0.077 |
| *Nausea\*\** | 1.04 | 0.27 | 0.79 | 1.36 | 0.786 |
| *Vomit\*\** | 0.68 | -1.33 | 0.39 | 1.20 | 0.183 |
| *Anosmia\** | 1.46 | 3.87 | 1.21 | 1.77 | <0.001 |
| *Dysgeusia\** | 1.36 | 3.12 | 1.12 | 1.64 | 0.002 |

**Figures S4-S5-S6.**
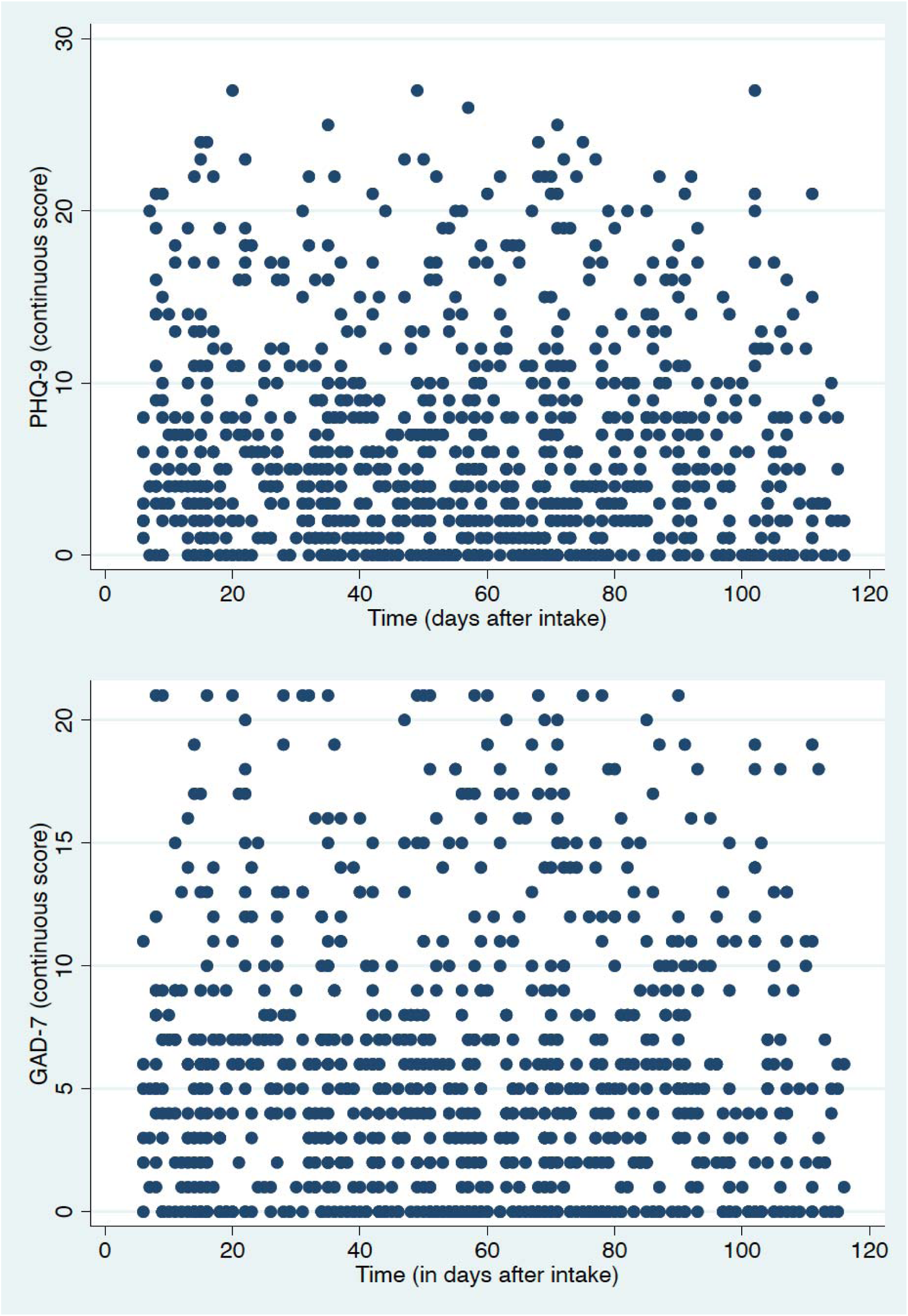
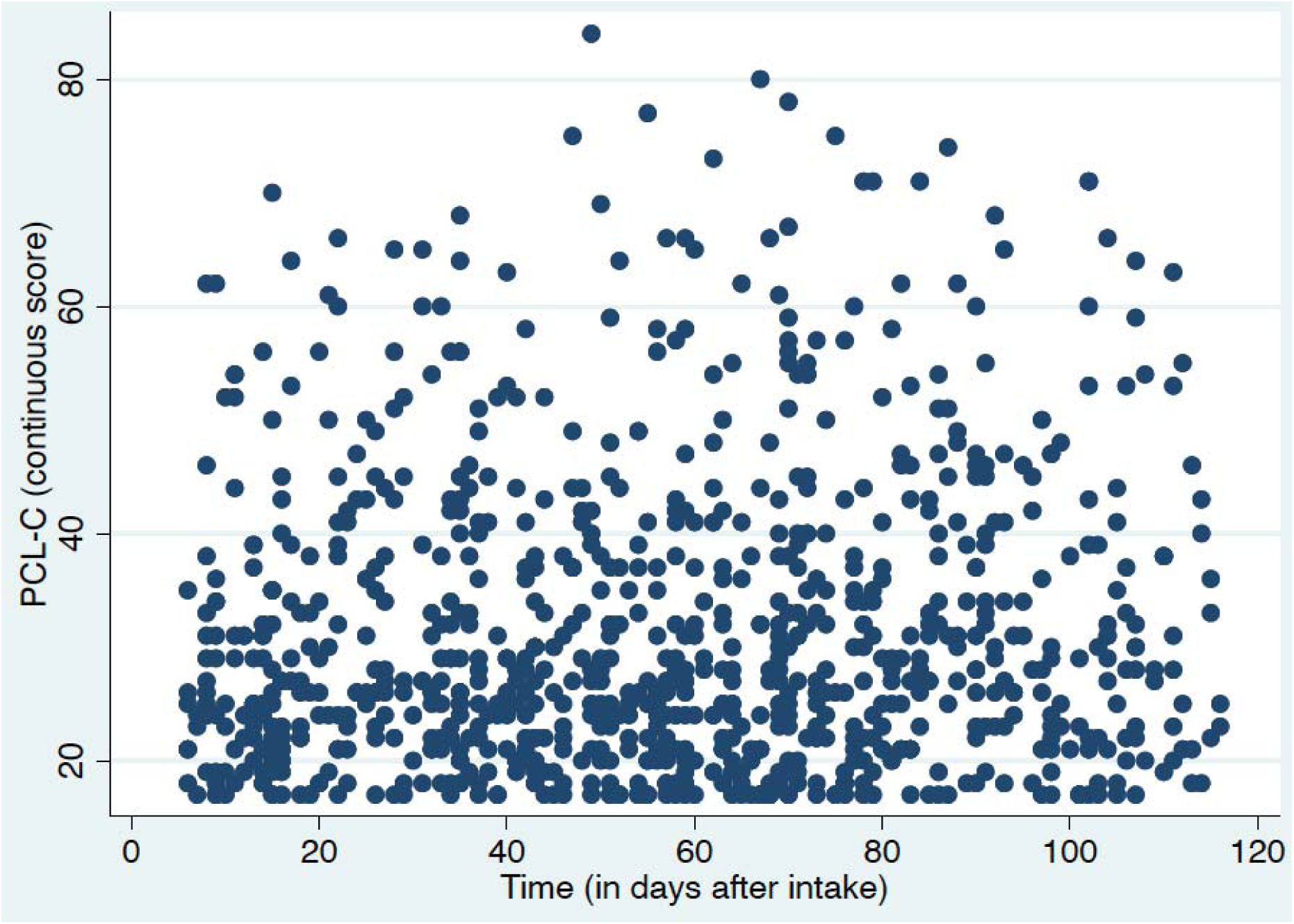
Scatterplots of scores of depression, anxiety, and post-traumatic stress (y-axis) by the time of the mental health assessment (x-axis).

## References

1. Kim MK, Jeon JH, Kim S, et al. The Clinical Characteristics and Outcomes of Patients with Moderate-to-Severe Coronavirus Disease 2019 Infection and Diabetes in Daegu, South Korea. Diabetes & Metabolism Journal 2020 [Advance online publication]. https://doi.org/10.4093/dmj.2020.0146

2. Aljabali A, Bakshi HA, Satija S, et al. COVID-19: Underpinning Research for Detection, Therapeutics, and Vaccines Development. Pharmaceutical Nanotechnology 2020. [Advance online publication].

3. Torales J, O’Higgins M, Castaldelli-Maia JM, Ventriglio A. The outbreak of COVID-19 coronavirus and its impact on global mental health. The International Journal of Social Psychiatry 2020; 66: 317–320.

4. Vindegaard N, Benros ME. COVID-19 pandemic and mental health consequences: Systematic review of the current evidence. Brain, Behavior, and Immunity 2020; S0889-1591(20)30954–5.

5. Lwin MO, Lu J, Sheldenkar A, et al. Global Sentiments Surrounding the COVID-19 Pandemic on Twitter: Analysis of Twitter Trends. JMIR public health and surveillance 2020; 6: e19447.

6. Fernández RS, Crivelli L, Guimet NM, Allegri RF, Pedreira ME. Psychological distress associated with COVID-19 quarantine: Latent profile analysis, outcome prediction and mediation analysis. Journal of affective disorders 2020; 27: 75–84.

7. Sinanović O, Muftić M, Sinanović S. COVID-19 Pandemia: Neuropsychiatric Comorbidity and Consequences. Psychiatria Danubina 2020; 33: 236–244.

8. Sparks SW. Posttraumatic Stress Syndrome: What Is It?. Journal of Trauma Nursing 2018; 25: 60–65.

9. Cheng SK, Tsang JS, Ku KH, Wong CW, Ng YK. Psychiatric complications in patients with severe acute respiratory syndrome (SARS) during the acute treatment phase: a series of 10 cases. The British Journal of Psychiatry 2004; 184: 359–360.

10. Wu KK, Chan SK, Ma TM. Post-traumatic stress, anxiety, and depression in survivors of severe acute respiratory syndrome (SARS). Journal of Traumatic Stress 2005; 18: 39–42.

11. Lee AM, Wong JG, McAlonan GM, et al. Stress and psychological distress among SARS survivors 1 year after the outbreak. Canadian journal of psychiatry 2007; 52: 233–240.

12. Guo Q, Zheng Y, Shi J, et al. Immediate psychological distress in quarantined patients with COVID-19 and its association with peripheral inflammation: A mixed-method study. Brain Behav Immun. 2020; 88: 17–27.

13. Hu, Y., Chen, Y., Zheng, Y., et al. Factors related to mental health of inpatients with COVID-19 in Wuhan, China, Brain Behav Immun. 2020 [Advanced online publication].

14. Nguyen HC, Nguyen MH, Do BN, et al. People with suspected COVID-19 symptoms were more likely depressed and had lower health-related quality of life: The potential benefit of health literacy. Journal of clinical medicine 2020; 9: 965.

15. Zhang J, Lu H, Zeng H, et al. The differential psychological distress of populations affected by the COVID-19 pandemic. Brain, behavior, and immunity 2020; 87: 49–50.

16. Leal FE, Mendes-Correa MC, Buss LF, et al. A primary care approach to the COVID-19 pandemic: clinical features and natural history of 2,073 suspected cases in the Corona São Caetano programme, São Paulo, Brazil. MedRvix. Doi: 10.1101/2020.06.23.20138081

17. Spitzer RL, Kroenke K, Williams JB, Löwe B. A brief measure for assessing generalized anxiety disorder: the GAD-7. Archives of internal medicine 2006; 166: 1092–1097.

18. Kroenke K, Spitzer RL, Williams JB, Monahan PO, Löwe B. Anxiety disorders in primary care: prevalence, impairment, comorbidity, and detection. Annals of internal medicine, 2007; 146: 317–325.

19. Moreno AL, DeSousa DA, Souza amflpd, et al. Factor structure, reliability, and item parameters of the Brazilian-Portuguese version of the GAD-7 questionnaire. Temas em Psicologia 2014; 24: 367–376.

20. Muñoz-Navarro R, Cano-Vindel A, Moriana JA, et al. Screening for generalized anxiety disorder in Spanish primary care centers with the GAD-7. Psychiatry Res. 2017;256:312–317.

21. Spitzer RL, Williams JB, Kroenke K, et al. Utility of a new procedure for diagnosing mental disorders in primary care. The PRIME-MD 1000 study. JAMA 1994; 272: 1749–1756.

22. Spitzer RL, Kroenke K, Williams JB. Validation and utility of a self-report version of PRIME-MD: the PHQ primary care study. Primary Care Evaluation of Mental Disorders. Patient Health Questionnaire. JAMA 1999; 282: 1737–1744.

23. Kroenke K, Spitzer RL, Williams JB. The PHQ-9: validity of a brief depression severity measure. J Gen Intern Med. 2001;16(9):606–613.

24. de Lima Osório F, Vilela Mendes A, Crippa JA, Loureiro SR. Study of the Discriminative Validity of the PHQ-9 and PHQ-2 in a Sample of Brazilian Women in the Context of Primary Health Care. Perspectives in psychiatric care 45: 216–227.

25. Levis B, Benedetti A, Thombs BD; DEPRESsion Screening Data (DEPRESSD) Collaboration. Accuracy of Patient Health Questionnaire-9 (PHQ-9) for screening to detect major depression: individual participant data meta-analysis. BMJ. 2019;365:1476.

26. Weathers FW, Litz BT, Herman DS, Huska JA, Keane TM. The PTSD Checklist (PCL): Reliability, validity, and diagnostic utility. In annual convention of the international society for traumatic stress studies. San Antonio, TX, Vol. 462, 1993.

27. Berger W, Mendlowicz MV, Souza W, Figueira I. Semantic equivalence of the Portuguese version of the Post-Traumatic Stress Disorder Checklist-Civilian Version (PCL-C) for the screening of post-traumatic stress disorder. Revista de Psiquiatria do Rio Grande do Sul 2004; 26: 167–175.

28. Lima EDP, Barreto SM, Assunção AÁ. Factor structure, internal consistency and reliability of the Posttraumatic Stress Disorder Checklist (PCL): an exploratory study. Trends in psychiatry and psychotherapy 2012; 34: 215–222.

29. Archer KR, Heins SE, Abraham CM, Obremskey WT, Wegener ST, Castillo RC. Clinical Significance of Pain at Hospital Discharge Following Traumatic Orthopedic Injury: General Health, Depression, and PTSD Outcomes at 1 Year. Clin J Pain. 2016;32(3):196–202.

30. Gustavsson S, Fagerberg B, Sallsten G, Andersson EM. Regression models for log-normal data: comparing different methods for quantifying the association between abdominal adiposity and biomarkers of inflammation and insulin resistance. International journal of environmental research and public health 2014; 11: 3521–3539.

31. Wang YX, Guo HT, D. XW, Song W, Lu C, Hao WN. Factors associated with post-traumatic stress disorder of nurses exposed to corona virus disease 2019 in China. Medicine 2020; 99: e20965–e20965.

32. Tang W, Hu T, Hu B, et al. Prevalence and correlates of PTSD and depressive symptoms one month after the outbreak of the COVID-19 epidemic in a sample of home-quarantined Chinese university students. Journal of affective disorders 2020 [Epub ahead of print]

33. Liang L, Gao T, Ren H, et al. Post-traumatic stress disorder and psychological distress in Chinese youths following the COVID-19 emergency. Journal of Health Psychology 2020; 25: 1164–1175.

34. González-Sanguino C, Ausín B, Ángel Castellanos M, al. Mental Health Consequences during the Initial Stage of the 2020 Coronavirus Pandemic (COVID-19) in Spain. Brain, Behavior, and Immunity 2020 [Epub ahead of print]

35. Lin CY, Peng YC, Wu YH, Chang J, Chan CH, Yang DY. The psychological effect of severe acute respiratory syndrome on emergency department staff. Emergency Medicine Journal 2007; 24: 12–17.

36. de Castro Longo MS, Vilete LMP, Figueira I, et al. Comorbidity in post-traumatic stress disorder: A population-based study from the two largest cities in Brazil. Journal of Affective Disorders 2020; 263: 715–721.

37. Park HY, Park WB, Lee SH, et al. Posttraumatic stress disorder and depression of survivors 12 months after the outbreak of Middle East respiratory syndrome in South Korea. BMC public health 2020; 20: 1–9.

38. Oosterbaan V, Covers M, Bicanic I, Huntjens R, de Jongh A. Do early interventions prevent PTSD? A systematic review and meta-analysis of the safety and efficacy of early interventions after sexual assault. European journal of psychotraumatology 2019; 10: 1682932.

39. Wei N, Huang BC, Lu SJ, et al. Efficacy of internet-based integrated intervention on depression and anxiety symptoms in patients with COVID-19. Journal of Zhejiang University. Science 2020; 21: 400–404.

40. Dubey S, Biswas P, Ghosh R, et al. Psychosocial impact of COVID-19. Diabetes & metabolic syndrome 2020; 14: 779–788. Advance online publication.

41. Iglesias-Sánchez PP, Vaccaro Witt GF, Cabrera FE, Jambrino-Maldonado C. The Contagion of Sentiments during the COVID-19 Pandemic Crisis: The Case of Isolation in Spain. International journal of environmental research and public health 2020; 17: E5918.

